# One-Year Clinical Outcomes of Biodegradable Polymer Coated Everolimus-Eluting Coronary Stent: Results From a Prospective, Open-Label, Non-Randomized Study

**DOI:** 10.1101/2024.12.16.24319090

**Authors:** Sridhar Kasturi, Shailender Singh, Vijay Kumar Reddy, Vikram Pratap, Abhishek Masalawala, Anil Kumar Mishra

## Abstract

**Purpose:** Newer-generation biodegradable polymer coated drug-eluting stents have been developed to optimize the outcomes of percutaneous coronary intervention. Everoshine everolimus eluting coronary stent (EECS) is a newer-generation, ultrathin (65 µm) biodegradable polymer–coated drug-eluting stent designed on a cobalt– chromium stent platform. This study aimed to evaluate safety and efficacy of the newer generation polymer-coated Everoshine EECS in patients with coronary artery disease (CAD) attributable to native coronary artery stenosis.

**Methods:** This was a single-center, single-arm, prospective study, which enrolled patients aged ≥ 18 years who had at least one native coronary artery lesions and were eligible for Everoshine EECS implantation. The primary endpoint was major adverse cardiac events (MACE), a composite of cardiac death, myocardial infarction (MI) and target lesion revascularization, assessed at the 30-day follow-up.

**Results:** A total of 193 patients were enrolled (mean age 57.8 ± 11.0 years; 142 (73.6%) male). Of these, 92 (47.7%) patients had ST-segment elevation MI (STEMI) and 39 (20.2%) had non-STEMI. A total of 267 stents were deployed, with a mean diameter of 2.9 ± 0.4 mm and a mean length of 27.0 ± 9.9 mm. At 30-day, 3 (1.6%) patients experienced MACE, and 2 (1.0%) patients experienced stent thrombosis. Cumulative MACE was observed in 5 (2.6%) patients at 1 year.

**Conclusion:** One-year clinical outcomes demonstrated satisfactory efficacy and safety of the novel biodegradable polymer-coated Everoshine EECS in patients with CAD attributable to native coronary artery stenosis.

The trial is registered retrospectively <trial registration number: ISRCTN13284341 dated 2 December 2024>.

## INTRODUCTION

Coronary artery disease (CAD) is a common cause of morbidity and mortality worldwide [1]. In India, CAD ranks among the top five causes of mortality. The CAD-related mortality is projected to rise rapidly, surpassing that in high-income countries [2].

Percutaneous coronary intervention (PCI) with stent implantation has revolutionized the treatment of CAD. Drug-eluting stents (DES) represent a significant advancement in PCI, radically transforming its practice. Data from the National Interventional Council 2016 registry showed that nearly 800 catheterization laboratories across India performed PCI with DES implantation in over 95% of the procedures [3].

First-generation durable polymer–coated DES reduce neointimal proliferation, thereby decreasing the risk of in-stent restenosis or the need for repeat revascularization compared with bare-metal stents. However, these first-generation durable polymer–coated DES have been associated with an increased incidence of stent thrombosis (ST) [4-7]. Although the occurrence of ST is rare, it results in mortality in up to 45% of cases and nonfatal myocardial infarction (MI) in another 30% – 40% of cases [8-10]. Several pathological and pre-clinical studies have demonstrated that the persistence presence of polymers is associated with prolonged inflammation of the coronary vessel wall, delayed arterial healing, and formation of neoatherosclerosis, which increases the risk of late (30 days to 1 year after stent implantation) and very late (more than 1 year after stent implantation) ST [11-13]. To overcome the challenges of first-generation durable polymer–coated DES, second-generation DES were developed with more biocompatible polymer coatings, improved antiproliferative drug selection and thinner metal alloy stent (cobalt–chromium or platinum–chromium) platforms. However, as the permanent polymer coating of second-generation DES remains a potential trigger for the occurrence of very late ST, newer-generation DES with biodegradable polymers were introduced to mitigate these limitations. It is hypothesized that biodegradable polymer coated DES provide controlled release of antiproliferative drug, thereby increasing the efficacy of DES by reducing in-stent restenosis while gradual degradation of the polymer coating offers a safety profile similar to that of bare-metal stents by lowering the incidence of ST [14]. Furthermore, the flexible designs and ultrathin strut thickness of the newer-generation biodegradable polymer–coated DES enhance stent apposition and accelerate strut endothelialization, leading to improved clinical outcomes [14]. Clinical studies and meta-analyses have shown superior clinical outcomes with newer-generation biodegradable polymer–coated DES, particularly in lowering the risk of very late ST (>1 year) and improving patient-orientated composite endpoint, including all-cause death, any MI, and all-cause or target vessel revascularization compared with durable polymer–coated DES [15-18].

The Everoshine (Kamal Encon Industries Limited, Faridabad, India) is a newer-generation biodegradable polymer–coated everolimus-eluting coronary stent (EECS) with ultrathin strut thickness. The Everoshine EECS has been approved by the Central Drugs Standard Control Organization (CDSCO) in 2019 for use in patients eligible for percutaneous transluminal coronary angioplasty. The Everoshine EECS is indicated for improving coronary luminal diameter in patients with symptomatic ischemic disease caused by discrete de novo lesions ≤ 47 mm in length in native coronary arteries with a reference vessel diameter of 2.00 – 4.50 mm. The stent has also received Conformité Européenne (CE) mark approval. This study aimed to evaluate the safety and efficacy of Everoshine EECS in patients with CAD attributable to native coronary artery stenosis.

## METHODS

### Study design

This prospective, open-label, non-randomized, single-arm study was conducted in adult patients (aged ≥ 18-years) with ≥ 1 coronary artery lesion at Sunshine Hospitals, Secunderabad, India, between September 2020 and August 2022.

### Patient population

Patients aged ≥ 18 years with at least one native coronary artery lesion and meeting eligibility criteria (indication, lesion length and diameter) for implantation of the study device were included in the study. The key exclusion criteria were as follows: 1) Patients with a native coronary artery lesion in which complete inflation of an angioplasty balloon was not feasible per the investigator’s discretion; 2) Patients with a known left ventricular ejection fraction < 30%; 3) Patient with a history of heart transplant or any other organ transplant, or those on the waiting list for any organ transplant; 4) Patients with known hypersensitivity or allergies to aspirin, heparin, clopidogrel, ticlopidine, everolimus or similar drugs or analogs/derivatives of everolimus, cobalt, chromium, nickel, molybdenum or contrast media; 5) Patients for whom antiplatelet and/or anticoagulation therapy was contraindicated; and 6) Patients receiving or expected to receive the chronic anticoagulation therapy (e.g., heparin or warfarin).

### Study device

The Everoshine EECS (Kamal Encon Industries Limited, Faridabad, India) is intended for use in patients with symptomatic ischemic disease caused by discrete de novo lesions ≤ 47 mm in length in native coronary arteries, with a reference vessel diameter of 2.00 – 4.50 mm. This next-generation variable geometry stent consists of an L605 cobalt-chromium alloy stent platform coated with a biodegradable polymer mixture, and everolimus as the active ingredient (**Figure 1**). This stent design features an open-cell design with uniform sequential rings throughout the stent. This design allows for unidirectional expansion in a controlled manner, ensuring uniform force distribution across the stent length and minimizing arterial injury. Furthermore, the stent design reduces the risk of edge flaring during expansion and minimizes recoil after balloon deflation, thereby preventing malposition and restenosis. The open-cell design also provides high radial strength at the edges, making it ideal for ostium stenting. The struts exhibit a wavy pattern, enhancing radial support (with an assigned number of crowns and connectors) and improving trackability performance. The strut thickness of the stent is 65 ± 10 µm, which minimizes focal mechanical injury to the endothelium and media. The biodegradable polymers mixture is a unique matrix of proven biocompatible, biodegradable polymers namely Poly L-Lactide, 50/50 Poly DL-Lactide-co-Glycolide. This mixture enables the programmed release of everolimus drug (drug load: 1.2 µg/mm^2^) for up to 48 days with drug elution completing within 3–7 weeks. The polymers are evenly coated on the stent surface, and the polymer chains are cleaved by hydrolysis to form monomeric acids, which eliminated from the body via urine; This complete degradation of polymers takes approximately 3–6 months. The Everoshine EECS is commercially available in lengths of 8, 9, 12, 13, 15, 16, 18, 20, 23, 24, 28, 32, 33, 36, 38, 40, 43, and 47 mm and available diameters of 2, 2.25 2.5, 2.75, 3.0, 3.5, 4 and 4.5 mm.

**Figure 1.**
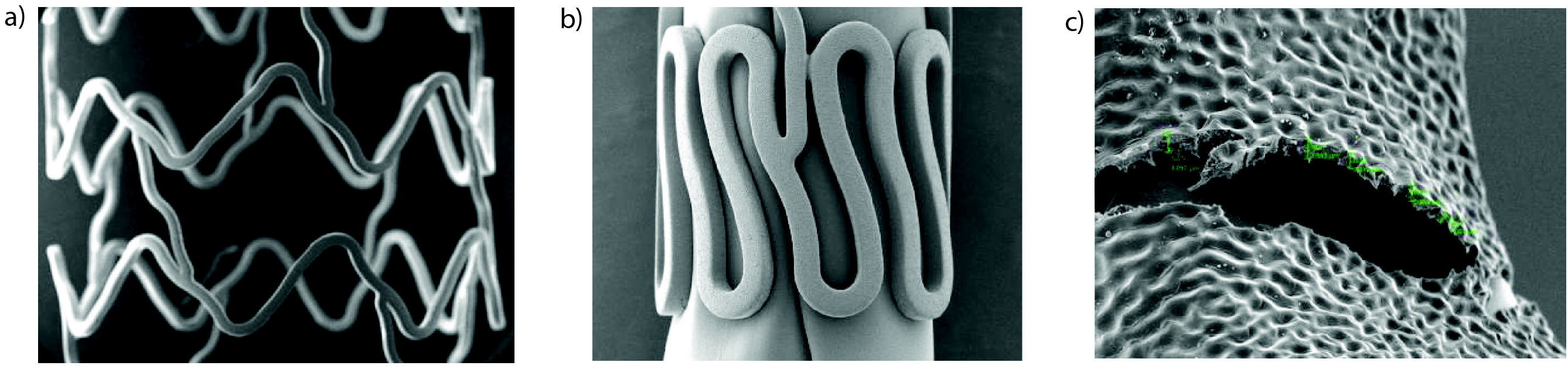
Scanning electron microscope images of (a) normal, (b) crimped and (c) expanded stent

### Interventional procedure and adjunctive medication

The procedure was performed according to institutional standard treatment guidelines. Briefly, either the femoral or radial artery was accessed using an introducer needle, followed by the placement of a sheath introducer to keep the artery open and control bleeding. A guiding catheter was advanced to the coronary artery opening and radiopaque dyes were injected through the guiding catheter into the coronary artery to perform coronary angiography and to determine the disease state and lesion size. Based on the angiographic findings, investigators determined the type of balloon catheter and coronary guidewire as well as the appropriate stent length/diameter. The coronary guidewire was inserted and navigated to the site of stenosis through the guiding catheter. A balloon catheter together with a deflated balloon was introduced and advanced to the blockage. Once inside the blockage, the balloon was inflated to compress the atheromatous plaque and expand the artery. Subsequently, the study device mounted on the balloon was introduced and expanded to complete the implantation (at the lesion location) in the culprit vessel.

All patients received a loading dose of aspirin (300 mg). Heparin or bivalirudin was used for procedural anticoagulation. The administration of a glycoprotein IIb/IIIa inhibitor during the procedure was at the investigator’s discretion. The patients received dual antiplatelet therapy (aspirin 75–300 mg/day indefinitely and either clopidogrel 75 mg/day, prasugrel 10 mg/day, or ticagrelor 90 mg twice daily for at least 12 months) after the procedure.

### Study follow-up

All patients were followed up at the clinical site at 30 days (± 7 days) and via telephone at 1 year (± 30 days).

At the 30-day and 1-year follow-up, post-procedural events, including MI, revascularization, ST, and death were evaluated. At 30-days, major bleeding complications (such as access site hematoma, retroperitoneal hemorrhage, intracranial hemorrhage, pericardial effusion/tamponade, or bleeding due to antiplatelet or anticoagulant therapy) and compliance to antiplatelet therapy were also evaluated.

### Study Endpoints

The primary endpoint was a major adverse cardiac event (MACE) at the 30-day follow-up. MACE was defined as the composite of cardiac death, MI and target lesion revascularization (TLR). The secondary endpoints included cardiac death, non-cardiac death, MI, and TLR at the 1-year follow-up. The occurrence of ST was also assessed at both timepoints, 30 days and 1 year.

Cardiac death was defined as death due to any cardiac cause (such as MI, low-output failure, or lethal arrhythmia), unwitnessed death, death of unknown cause, or all procedure-related deaths linked to concomitant treatment. Noncardiac death was defined as death with a well-established noncardiac cause. MI was considered if troponin values were elevated greater than five times upper normal limit (> 5 × 99^th^ percentile upper reference limit [URL]) in patients with normal baseline values (≤ 99^th^ percentile URL) or if troponins increased > 20% if baseline values were elevated and stable or falling. TLR was defined as any repeat revascularization (either PCI or coronary artery bypass grafting) of the lesion anywhere within the stent or within the subsequent 5 mm of distal or proximal segment to the stent. ST was defined per the Academic Research Consortium criteria [19].

### Statistical analysis

Data were analyzed using descriptive statistics. Continuous variables are presented as mean ± standard deviation, and categorical variables as counts and percentages. All data were analyzed using Microsoft Office Excel.

## RESULTS

A total of 193 patients were screened and enrolled in the study. Baseline demographics and clinical characteristics of patients are summarized in **Table 1**. The average age of the patients was 57.8 ± 11.0 years, with the majority being male (n = 142; 73.6%). Hypertension (n = 90; 46.6%) and diabetes (n = 68; 35.2%) were the most common coronary risk factors. The history of PCI was noted in 12 (6.2%) patients. The average left ventricular ejection fraction was 50.7 ± 9.1%. The majority of the patients were diagnosed with MI, with 92 (47.7%) having STEMI and 39 (20.2%) having NSTEMI diagnosis.

**Table 1.** Baseline demographic characteristics.

| Characteristics | Number of patients (N = 193) |
| --- | --- |
| Age (years), mean ± SD | 57.8 ± 11.0 |
| Male sex, n (%) | 142 (73.6) |
| Alcoholics, n (%) | 8 (4.2) |
| Smokers, n (%) | 17 (8.8) |
| Body mass index (kg/m<sup>2</sup>), mean ± SD | 24.8 ± 4.1 |
| Cardiovascular risk factors, n (%) |  |
| Hypertension | 90 (46.6) |
| Diabetes | 68 (35.2) |
| Anemia | 30 (15.5) |
| Dyslipidemia | 44 (22.8) |
| Previous PCI | 12 (6.2) |
| Heart failure per NYHA classification, n (%) |  |
| Class I | 1 (0.5) |
| Class II | 192 (99.5) |
| Severity of angina as per CCS classification, n (%) |  |
| Class I | 192 (99.5) |
| Class II | 1 (0.5) |
| Family history of coronary artery diseases, n (%) | 9 (4.7) |
| Clinical presentation, n (%) |  |
| STEMI | 92 (47.7) |
| NSTEMI | 39 (20.2) |
| Unstable angina | 56 (29.0) |
| Chronic stable angina | 6 (3.1) |
| Left ventricular ejection fraction (%), mean ± SD | 50.68 ± 9.08 |
| Blood pressure |  |
| SBP (mm Hg), mean ± SD | 121.55 ± 15.93 |
| DBP (mm Hg), mean ± SD | 77.30 ± 10.74 |
| Laboratory investigations |  |
| Triglycerides (mg/dl), mean ± SD | 175.5 ± 98.1 |
| Troponin I (ng/ml), mean ± SD | 27.6 ± 44.8 |

CABG, coronary artery bypass grafting; CCS, Canadian Cardiovascular Society; DBP, diastolic blood pressure; LVEF, left ventricular ejection fraction; NSTEMI, non-ST-segment elevated myocardial infarction; NYHA, New
York Heart Association; PCI, percutaneous coronary intervention; SBP, systolic blood pressure; SD, standard deviation STEMI, ST-segment elevation myocardial infarction.

### Lesions and procedural characteristics

Coronary angiography demonstrated single-vessel disease in 96 (49.7%) patients and the majority of them were in the left anterior descending artery (n=139; 52.1%). Fifty-six (29.0%) patients had long lesions, and 29 (15.0%) had totally occluded lesions. The lesion and procedural characteristics are summarized in **Table 2**. A total of 267 stents were implanted, with an average stent length and diameter of 27.0 ± 9.9 mm and 2.9 ± 0.4 mm, respectively (**Table 2**). In the majority of patients (n=149; 77.2%), the radial artery was accessed to perform PCI. Predilatation was performed in 63 (32.6%) patients. During the procedure, three patients experienced dissection which was successfully treated. All patients (100%) achieved Thrombolysis in MI (TIMI)-3 flow after procedure (**Online Resource 1**).

**Table 2.** Lesion and procedural characteristics.

| Characteristics | Number of patients (N = 193) |
| --- | --- |
| Extent of coronary artery disease, n (%) |  |
| Single vessel disease | 97 (49.7) |
| Double vessel disease | 75 (38.9) |
| Triple vessel disease | 21 (10.9) |
| Patients with long lesions, n (%) | 56 (29.0) |
| Patients with totally occluded lesions, n (%) | 29 (15.0) |
| Lesion location, n (%) |  |
| Left anterior descending artery | 139 (52.0) |
| Right coronary artery | 79 (29.6) |
| Left circumflex artery | 48 (18.0) |
| Left main artery | 1 (0.4) |
| Type of lesions, n (%) |  |
| Calcific | 3 (1.6) |
| Fibrocalcific | 45 (23.3) |
| Fibrotic | 68 (35.2) |
| Lipidic | 66 (25.2) |
| Thrombotic | 49 (25.4) |
| Fibrolipid | 36 (18.7) |
| Total number of stents implanted | 267 |
| Number of implanted stents, n (%) |  |
| 1 | 125 (64.8) |
| 2 | 61 (31.6) |
| 3 | 7 (3.6) |
| Stent diameter, mean ± SD | 2.9 ± 0.4 |
| Stent length, mean ± SD | 27.0 ± 9.9 |
| Patients with overlapping stents, n (%) | 44 (22.79) |

SD, standard deviation.

### Clinical outcomes

All patients (n=193) completed on-site clinical and telephonic follow-up at 30 days and 1 year, respectively. Except for one patient, all the patients were compliant with antiplatelet therapy at the 30-day follow-up.

At 30 days, 3 (1.6%) patients experienced MACE: 1 (0.5%) cardiac death, 2 (1.0%) MIs and 2 (1.0%) TLRs (**Table 3**). At 1 year, cumulative MACE occurred in 5 (2.6%) patients, which included 3 (1.6%) cardiac deaths, 2 (1.0%) MIs and 2 (1.0%) TLRs. At 30 -days, 2 (1.0%) patients experienced ST (**Table 3**). There were no occurrences of ST after 30 day until the 1-year follow-up. There were 2 noncardiac deaths.

**Table 3.** Cumulative clinical outcomes at the 30-day and 1-year follow-up.

| **Clinical outcomes** | **30-day follow-up**<br>**(N = 193 patients)** | **1-year follow-up**<br>**(N = 193 patients)** |
| --- | --- | --- |
| All-cause death, n (%) | 3 (1.6) | 5 (2.6) |
| Cardiac | 1 (0.5) | 3 (1.6) |
| Non-cardiac | 2 (1.0) | 2 (1.0) |
| MI, n (%) | 2 (1.0) | 2 (1.0) |
| TLR, n (%) | 2 (1.0) | 2 (1.0) |
| ST, n (%) | 2 (1.0) | 2 (1.0) |
| MACE,<sup>a</sup> n (%) | 3 (1.6) | 5 (2.6) |

<sup>a</sup>MACE was the composite of cardiac death, MI, and TLR
MACE, major adverse cardiac events; MI, myocardial infraction; ST, stent thrombosis; TLR, target lesion revascularization.

No major Bleeding complications were reported at the 30-day follow-up. However, 3 (1.15%) patients experienced hemorrhagic stroke at 30 days, all of which were successfully treated.

## DISCUSSION

This study assessing the safety and efficacy of the newer-generation Everoshine biodegradable polymer–coated EECS demonstrated that EECS was associated with low rates of MACE and overall ST at 30 days with these rates remaining consistently low at 1 year.

The technology of DES has evolved significantly since its inception. Although first-generation durable polymer–coated DES reduced the risk of restenosis compared with bare-metal stents, they were associated with an increased risk of late thrombotic events. The durable polymer coating of DES leads to a persistent inflammatory reaction in the vessel wall and can trigger hypersensitivity reactions, leading to incomplete strut endothelialization, thereby resulting in late thrombotic events [12, 20]. These challenges were addressed with the development of newer-generation biodegradable polymer–coated DES. Several clinical studies and meta-analyses have confirmed that biodegradable polymer–coated DES are superior to the first-generation durable polymer–coated DES [15-18]. Furthermore, strut thickness in DES impacts on stent thrombogenicity and restenosis during the early phase of stent implantation [21]. Kastrati et al., in a multicenter randomized trial (ISAR-STEREO trial) evaluated the association of angiographic and clinical restenosis with thin-strut stents (strut thickness 50 μm) and thick-strut stents (strut thickness 140 μm) in 651 patients with CAD. They found that angiographic restenosis (15.0% vs 25.8%) and repeat revascularization (8.6% vs 13.8%) were significantly lowered in patients treated with thin-struts stent than those treated with thick-strut stents [22]. Similar findings were reported in the ISAR-STEREO 2 trial, which compared thick-strut and thin-strut stents with different stent designs [23]. Considering the clinical benefits of low strut thickness, newer-generation DES have been developed with ultrathin strut [24]. The improved clinical outcomes observed in the present study may be attributed to the combination of ultrathin struts and biodegradable polymer coating EECS.

The present study intentionally included a wide range of the patients with CAD so that the enrolled patients truly represent the typical patient population encountered in a catheterization laboratory in routine clinical practice in India. Of all the patients enrolled in the study, 67.9% had MI, 35.2% had diabetes, and a considerable proportion had difficult-to-treat lesions (totally occluded lesions and long lesions) or multivessel disease. Despite the complexity of the enrolled patients’ clinical profiles in this study, low rates of MACE (2.6%) and ST (1.0%) were reported, which are consistent with those observed in previous studies of newer-generation biodegradable polymer–coated DES [25-27]. However, a head-to-head comparison of MACE and ST rates should be made with caution, owing to the differences in study populations, MACE definitions, and clinical follow-up.

### Limitations

This study has some limitations that should be considered when interpreting the results. The study had a single-arm, non-randomized design, which limits involvement of the comparison arm. Hence, the study lacks direct a head-to-head comparison with other DES to confirm the efficacy and safety of biodegradable polymer–based EECS. Moreover, the 1-year clinical follow-up was conducted via telephone, which may have led to underreporting of some events by the patients. However, most of the underreported MACE were observed to be asymptomatic events that were usually associated with less clinical significance than symptomatic events, suggesting that underreporting of MACE in the present study likely had limited impact on the study conclusions.

## Conclusion

The results of the present study demonstrated satisfactory efficacy and safety of the novel biodegradable polymer–coated EECS for treatment of CAD, with low rates of MACE and ST at the 1-year follow-up. Long-term comparative trials may offer future prospects for a head-to-head comparison of the efficacy of the EECS with other DES.

## Data Availability

The data that support the findings of this study are available from the corresponding author upon reasonable request.

## Declarations

## Acknowledgements

Medical writing assistance was provided by Iwana Consultancy Solution Private Limited, which was funded by Kamal Encon Industries Limited, Faridabad, India.

## Ethics approval

The study was conducted in accordance with the Declaration of Helsinki, ISO 14155 and country-specific regulatory requirements (ISRCTN13284341; the trial was registered retrospectively).

Institutional Ethics Committee of Sunshine Hospital gave approval for this work.

## Consent to participate

All patients provided written informed consent, which was reviewed and approved by the Institutional Ethics Committee.

## Author contribution

All authors were involved in drafting the article and revising it critically for important intellectual content, and all authors approved the final version to be published.

Sridhar Kasturi had full access to all the data in the study and takes responsibility for the integrity of the data and the accuracy of the data analysis.

Study conception and design: Sridhar Kasturi and Abhishek Masalawala.

Acquisition of data: Vijay Kumar Reddy and Shailendra Singh.

Analysis and interpretation of data: Sridhar Kasturi, Shailender Singh, Vijay Kumar Reddy, Vikram Pratap, Abhishek Masalawala, Anil Kumar Mishra.

## Supplementary Material

**Online Resource 1** Procedural details and complications

