## Supplementary material for "One-Year Clinical Outcomes of Biodegradable Polymer Coated Everolimus-Eluting Coronary Stent: Results From a Prospective, Open-Label, Non-Randomized Study": Online material

**Online Resource 1:** Procedural details and complications

| **Characteristics** | **Number of patients = 193** |
| --- | --- |
| Procedural medications, n (%) | |
| Aspirin | 193 (100) |
| Ticagrelor | 193 (100) |
| Glycoprotein IIb/IIIa inhibitorsblockers | 1 (0.5) |
| Heparin | 193 (100) |
| Statins | 193 (100) |
| Intracoronary nitroglycerine | 6 (3.1) |
| Intracoronary nicorandil | 5 (2.6) |
| Procedural details | |
| Route for PCI, n (%) |  |
| Radial artery | 149 (77.2) |
| Femoral artery | 43 (22.3) |
| Predilatation, n (%) | 63 (32.6) |
| Pre-procedural TIMI flow, n (%) |  |
| TIMI-1 | 33 (17.1) |
| TIMI-2 | 68 (35.2) |
| TIMI-3 | 166 (86.0) |
| Post-procedural TIMI flow, n (%) |  |
| TIMI-3 | 193 (100) |
| Procedural complications, n (%) |  |
| Dissection | 3 (1.6) |
| Slow flow | 1 (0.5) |

PCI, percutaneous coronary intervention; TIMI, Thrombolysis in Myocardial Infarction.
